# Feasibility of a community-based intervention for the diagnosis and management of hypertension in two rural populations in Kenya and The Gambia: IMPLEMENT-IHCoR Feasibility Study Protocol

**DOI:** 10.64898/2026.01.20.26344202

**Authors:** Syreen Hassan, Nancy Kagwanja, Brahima Diallo, Ruth Willis, Jasmine Hine, Aurelia Brazeal, Vallery Obure, Catherine Kalu, Clement Mwagwabi, Anoop Shah, Noni Mumba, Robinson Oyando, Alexander Perkins, Ellen Nolte, Benjamin Tsofa, Edwine Barasa, Pablo Perel, Modou Jobe, Anthony O. Etyang, Adrianna Murphy

**Affiliations:** Department of Noncommunicable Disease Epidemiology, Faculty of Epidemiology and Population Health, London School of Hygiene and Tropical Medicine, London, United Kingdom; Department of Health Systems and Research Ethics, Kenya Medical Research Institute Wellcome Trust Research Programme (KEMRI-WTRP), Kilifi, Kenya; Department of Nutrition and Planetary Health, Medical Research Council Unit The Gambia (MRCG) at London School of Hygiene and Tropical Medicine, Banjul, The Gambia; Department of Health Services Research and Policy, Faculty of Public Health and Policy, London School of Hygiene and Tropical Medicine, London, United Kingdom; Department of Epidemiology and Demography, KEMRI Wellcome Trust Research Programme, Kilifi, Kenya; Health Economics Research Unit, KEMRI Wellcome Trust Research Programme, Nairobi, Kenya

**Keywords:** Protocol, feasibility, hypertension, implementation, Kenya, Gambia

## Abstract

**Introduction:** Hypertension is the leading global risk factor for mortality, causing over 10 million deaths annually. In sub-Saharan Africa, hypertension prevalence is high, particularly in rural areas, where it is less likely to be diagnosed, treated, or controlled effectively. This results in a high burden of complications, including heart failure, stroke, and kidney disease. Community-centred approaches using community health workers (CHWs), risk-based approaches and simplified treatment regimens have shown promise in improving hypertension management. However, there is limited evidence on the effectiveness of such approaches in rural sub-Saharan Africa. This study aims to evaluate the feasibility and acceptability of a community-based intervention to improve hypertension management in rural Kenya and The Gambia.

The primary aim of this study is to assess the feasibility of a community-centred intervention for hypertension management in rural Kenya and The Gambia. The objectives are to evaluate the intervention’s adoption, fidelity, reach, and dose; understand the mechanisms of action and contextual factors affecting its implementation; assess acceptability from the perspectives of patients, healthcare providers, and policymakers; estimate the costs associated with the intervention; and evaluate study procedures to inform the design of a future full-scale trial.

**Methods and Analysis:** We will conduct a mixed-methods, non-randomised, single-arm feasibility study, designed in accordance with the CONSORT framework and checklist for feasibility and pilot studies, including best practice guidance for non-randomised feasibility studies. The study will be conducted in two rural sites: Kilifi, Kenya, and Kiang West, The Gambia.

The intervention was co-designed with stakeholders and includes community-based hypertension screening by CHWs, risk stratification, and hypertension-mediated organ damage (HMOD) assessment at primary healthcare facilities, followed by treatment initiation using single-pill combination (SPC) antihypertensive therapy for eligible individuals. Training will be provided to all healthcare providers involved in the study. We will screen 500 participants aged 30-80 years at their residence (250 from each country) and we expect that about 45% will be referred for additional assessments and of these 25% (or 10% of the total sample) will be prescribed treatment with SPC. Data collection to evaluate the intervention and its implementation will involve quantitative measures of feasibility and clinical outcomes; observations to assess fidelity and costing measures; and qualitative interviews and focus group discussions with patients, healthcare providers, and policymakers to understand the acceptability and contextual influences on intervention implementation.

**Ethics and dissemination:** Ethics approval was obtained from the Kenyan National Committee for Science, Technology and Innovation (NACOSTI) (ref: 415561), the Gambia Government/Medical Research Council Joint Ethics Committee (ref: 31372) and the London School of Hygiene and Tropical Medicine Ethics Committee (ref: 31372). Study findings will be disseminated through peer-reviewed publications, conferences, policy briefs, community engagement forums, and accessible summaries shared via the Improving Hypertension Control in Rural sub-Saharan Africa (IHCoR-Africa) and partner newsletters.

**Trial registration:** This study is registered with the ISRCTN- The UK’s Clinical Study Registry ( ), and Pan African Clinical Trials Registry (PACTR202504839027548).

**ARTICLE SUMMARY:** Strengths and limitations of this study

- This study uses a robust mixed-methods design incorporating both quantitative and qualitative data to evaluate feasibility, acceptability, and implementation of a complex intervention.
- The intervention was based on extensive formative health systems research, co-designed with stakeholders in two countries, and embedded within existing community and health system structures, enhancing its relevance and potential scalability.
- We are conducting a comprehensive training and using standardised protocols to ensure consistency in intervention delivery and data collection across study sites.
- As a single-arm feasibility study without a control group, findings on intervention effectiveness will be limited and primarily focused on informing a future definitive trial.

## INTRODUCTION

Hypertension is the leading risk factor for premature mortality globally, accounting for 10.8 million deaths annually [1, 2]. Sub-Saharan Africa (SSA) has among the highest hypertension prevalence worldwide, with nearly 700,000 deaths attributed to hypertension in 2019 — double the number in 1990 [2]. In SSA, hypertension tends to occur at younger ages, is more severe, and is poorly controlled, contributing to high rates of complications like heart failure, stroke, kidney disease, and premature death [3, 4]. A systematic review of 33 SSA studies found that only 27% of people with hypertension were aware of their status, 18% were receiving treatment, and just 7% had controlled blood pressure [5]. Rural areas are of particular concern, given weak health systems, low health literacy, and high cardiovascular case-fatality rates [6].

With 60% of SSA’s 1.1 billion people living in rural areas, hypertension prevalence is similar to urban areas, but awareness, treatment, and control are significantly lower [5–7]. For instance, Kenya’s 2015 STEPS survey found hypertension awareness in rural areas was only 12.3%, compared to 20% in urban settings [8]. To meaningfully reduce hypertension burden, improvements are needed across the full care cascade, from awareness and diagnosis to treatment, adherence, and monitoring [9]. Barriers in rural SSA are multifaceted. At the individual level, these include the asymptomatic nature of hypertension, low risk perception, caregiving duties, financial constraints, low adherence, and misconceptions about medication [10–13]. Provider-level barriers include inadequate training, poor communication, limited infrastructure, and weak referral systems [14]. At the system level, challenges include limited access to care, medication stockouts, unaffordable treatments, and chronic underinvestment in health services [15].

There is growing evidence from low- and middle-income countries where access to medical care is limited, that community-centred approaches (involving community health workers (CHWs), risk stratification, and simplified treatment using single-pill combination (SPC) therapy (2–3 blood pressure-lowering medications in one pill) can play a key role in managing hypertension and its consequences [15, 16]. The HOPE-4 trial (Colombia and Malaysia) and the COBRA study (South Asia) both demonstrated that CHW-led interventions, supported by electronic tools and simplified treatment protocols, significantly improved blood pressure control [17, 18]. These findings have informed global recommendations, including by the Pan-African Society of Cardiology (PASCAR) [19]and WHO’s HEARTS Technical Package [20], and are reinforced by systematic reviews showing that CHW-led interventions enhance linkage to care, treatment adherence, and blood pressure control in low- and middle-income countries (LMICs) [16, 21].

However, there is limited evidence on the effectiveness and implementation of such approaches in rural SSA. Key gaps remain regarding the appropriateness of risk-based screening and diagnosis methods, and the contextual factors that shape the feasibility and sustainability of simplified treatment approaches like SPCs [22]. From a public health perspective, the sustainability of community-centred hypertension programmes depends on a deep understanding of the broader context, including existing policies, organisational and institutional structures, and the experiences of those using (patients and family carers), delivering (healthcare workers), and overseeing services (decision-makers at different system levels).

To address these gaps, our research aims to evaluate the feasibility of a contextually adapted, community-based hypertension intervention. The intervention design was informed by work undertaken within two linked programmes: the Improving Hypertension Control in Rural sub-Saharan Africa (IHCoR-Africa) and the Improving Implementation of Innovations in CVD (IMPLEMENT-CVD) studies. IHCoR-Africa involved (1) understanding experiences and practices across patient, provider, and system levels; (2) determining optimal diagnostic and risk stratification methods; and (3) structured participatory action research workshops held in both countries to developing a community-centred hypertension management programme [23, 24]. The IMPLEMENT-CVD study focuses on the implementation of fixed-dose combination therapy for hypertension in Kenya. Work conducted between April 2022–April 2024 explored i) the acceptability of SPC therapy through qualitative interviews with patients, caregivers, and healthcare workers in Kiambu County [25], followed by consultations in Kilifi County to adapt findings to the local context and ii) the health systems barriers to implementation of FDCs in Kenya through interviews with policymakers [26].

This protocol outlines the feasibility study to assess the implementation of a co-designed, community-based hypertension intervention in rural Kenya and The Gambia.

## AIM AND OBJECTIVES

We aim to evaluate the feasibility and acceptability of a community-based intervention to improve hypertension management and treatment in rural Kenya and The Gambia. To achieve this, we have five specific objectives:

**Objective 1:** Evaluate the feasibility of the intervention in terms of adoption, fidelity, reach, and dose.

**Objective 2:** Understand the mechanisms of action and contextual factors affecting the feasibility of each intervention component.

**Objective 3:** Understand the acceptability and sustainability of the intervention from the perspective of patients, healthcare providers, research staff and policy makers.

**Objective 4:** Estimate the costs associated with the implementation of the intervention.

**Objective 5:** Evaluate the study procedures to inform the design of a future full-scale trial.

## TRIAL DESIGN

The IMPLEMENT-IHCoR study is a mixed-methods, non-randomised, single-arm feasibility study evaluating a novel hypertension management intervention in rural SSA. Conducted in Kenya and The Gambia, the study assesses the intervention’s feasibility and informs the design of a future definitive trial, with no predetermined progression criteria.

The study follows the CONSORT framework and checklist for feasibility and pilot studies, alongside best practice guidelines for non-randomised feasibility studies [27, 28]. It is also informed by the UK Medical Research Council Complex Intervention Framework and the Template for Intervention Description and Replication (TIDiER) checklist for developing and evaluating complex interventions [29, 30]. This protocol has been developed and reported in accordance with the SPIRIT (Standard Protocol Items: Recommendations for Interventional Trials) 2013 guidelines [31]. Qualitative evaluation will be guided by two complementary theoretical frameworks: the Theoretical Framework of Acceptability [32], assessing the intervention’s acceptability among deliverers and recipients, and Normalisation Process Theory, which examines the mechanisms affecting intervention integration into practice [33]. Together, these frameworks guide a comprehensive assessment at the individual and broader health system levels.

## METHODS

### Study sites

The study will be conducted in two sites in Kilifi County, Kenya, and three sites in Kiang West, The Gambia. Both regions have high hypertension prevalence and established research infrastructures including Health and Demographic Surveillance Systems (HDSS), from which a list of patients will be generated for recruitment.

Kilifi County in Kenya was selected for its well-developed research infrastructure and is one of Kenya’s poorest regions. The Kilifi Health and Demographic Surveillance System (KHDSS) covers part of the county (900 km²) with a population of approximately 300,000 [34]. The prevalence of hypertension is 26% and only 3% of individuals have their blood pressure controlled [35]. The selected rural sites for the study are Matsangoni (primarily from the Mijikenda ethnic group, predominantly Giriama) and Chasimba (primarily from the Mijikenda ethnic group, predominantly Chonyi). Community Health Promoters (CHPs), supervised by Community Health Assistants (CHAs) based at primary healthcare facilities, deliver preventive health services in defined geographic areas (Community Health Units or CHUs), each covering about 5,000 people. This study includes two rural CHUs: Matsangoni Health Centre (2 clinical officers, 1022 households, 12 CHPs), and Chasimba Health Centre (1 clinical officer, 1200 households, 26 CHPs). Recent national initiatives emphasise community-level screening for NCDs [36], including hypertension, with national and county governments collaboratively providing CHPs monthly stipends. Despite policy support, CHPs in the study sites have yet to commence routine hypertension screening.

Kiang West district in The Gambia covers 750 km², encompassing 36 villages and a population exceeding 14,000, with most residents classified in the lowest wealth groups In our analysis of the 2010 WHO STEPS NCD data, we found a hypertension prevalence in this region of 40%, of whom 71% were previously undiagnosed, and only 4% were controlled [37]. The region’s health service delivery is managed by the Regional Health Team in the Lower River Region. The selected rural villages for the study are Batelling (350 residents, primarily Mandinka, subsistence farming, 18 km from Jifarong Community Clinic), Kemoto (460 residents, predominantly Mandinka, subsistence farming and fishing, 10 km from Karantaba Health Centre), and Tankular (900 residents, predominantly Mandinka, subsistence fishing and farming, 17 km from Karantaba Health Centre). Four Community Health Nurses (CHNs), each overseeing clusters of 7-9 villages, supervise Village Health Workers (VHWs). There is one VHW in each village and they are selected by their respective communities to conduct health related activities on voluntarily basis and have no prior involvement in hypertension care services delivery in routine practice.

### Eligibility Criteria

The IMPLEMENT-IHCoR study will include participants from five communities, two in rural Kenya and three in rural Gambia. The eligibility criteria have been set broadly to reflect as closely as possible the general adult population of the study communities. Eligibility will be assessed in the community by trained CHPs (Kenya) and CHNs/VHWs (The Gambia) using the pre-generated lists from respective HDSS. Ineligible individuals will be replaced with matched alternatives. Eligible participants will be approached for informed consent by the same health workers.

#### Inclusion criteria

Participants must be aged 30 to 80 years at the time of consent. Individuals below 30 have low risk of CVD, while individuals over 80 may require additional safety considerations due to frailty. Participants must also be able to complete all study procedures.

#### Exclusion criteria

Individuals with self-reported pre-existing hypertension on current treatment will be excluded from the study. Pregnant and breastfeeding women, identified through self-report, will also be excluded, as their hypertension management is beyond the scope of this intervention.

### Recruitment

Participants aged 30–80 years will be randomly selected from the HDSS platforms in each study setting using age- and sex-stratified sampling. Age will be grouped into five strata (30–39, 40–49, 50–59, 60–69, 70–80), and sex stratified as male or female.

Replacement participants matched by strata and location will be included to account for ineligibility, loss to follow-up, or refusal.

### Patient and public involvement

Community engagement has been central to this programme from its inception. Patients and the public were first involved during the early planning stages of the programme through scheduled stakeholder meetings in both Kenya and The Gambia. In addition, this intervention’s design was informed by structured participatory action research workshops held in both countries. In The Gambia, three workshops held between May and August 2024 brought together 37 participants, including hypertensive patients, caregivers, village health workers, healthcare providers, managers, and NCD policy partners. In Kenya, three workshops held over the same period engaged 93 participants, including patients, carers, CHPs, CHAs, facility and county health managers, frontline providers, and MoH representatives. These workshops shaped the design and informed priorities, feasibility, and acceptability of intervention components.

While a formal assessment of participant burden was not conducted, participant and community insights gathered through workshops directly influenced the delivery model, ensuring that intervention activities would be brief, home-based, and minimally disruptive. Additionally, embedding data collection within existing community health structures through CHPs (Kenya) and CHNs/VHWs (The Gambia) supports integration and acceptability.

## INTERVENTION COMPONENTS AND TIMELINES

The study intervention will cover three areas: (a) task-sharing between clinicians and CHPs or VHWs for blood pressure screening, monitoring, and adherence follow-up; (b) diagnosis and risk stratification based on HMOD assessment and history of CVD at primary healthcare facilities; and (c) dispensing SPC medication at initial diagnosis by clinicians, with subsequent community-level refills. We will use Perindopril + Amlodipine (an ACE inhibitor + calcium channel blocker), based on availability and evidence of its suitability for Black African populations [38]. For women of childbearing age (30-49 years), we will use Amlodipine (5 mg) + Hydrochlorothiazide (12.5 mg). For those not achieving control on dual therapy, we will add hydrochlorothiazide (a diuretic) which aligns with international and Kenyan guidelines [39].

Community screening by CHPs/VHWs involves attended Automated Home Blood Pressure measurement (aAHBP) following international recommendations. The CHP/VHW will use an automated blood pressure machine (Omron M7 Intelli IT) to collect three measurements of the participant’s blood pressure in a seated position. An average of the three measurements will be calculated by the machine and the decision to refer to a primary healthcare facility will be based on this average. Participants with normal blood pressure £130/80 will have a final follow-up visit at six months.

Participants with blood pressure ≥130/80mmHg will be referred to primary healthcare facilities for further assessment. Those with blood pressure ≥180/110mmHg will be referred to a higher clinical level for expedited review. If those participants are showing hypertensive crisis symptoms, they will be excluded from the study, and if asymptomatic, initiated on SPC and retained.

At primary healthcare facilities, clinicians will perform an attended Automated Office Blood Pressure measurement (aAOBP), assess HMOD via point-of-care tests (ECG, urine dipstick), and review CVD risk factors. Participants will then be classified for treatment initiation of SPC hypertension treatment and/or life-style advice based on the following:

Participants with blood pressure <160/108mmHg without diabetes, previous CVD, or HMOD will receive lifestyle advice only. These participants will have a final follow-up visit from the CHPs/VHWs at six months after the initial clinic visit. Participants with blood pressure ≥130/80mmHg with diabetes, previous CVD, or HMOD or those with blood pressure > 160/100mmHg, will be initiated on SPC.

Participants initiated on SPC therapy will be followed up regularly over six months by CHPs in Kenya and VHWs/CHNs in The Gambia. Follow-up visits will focus on measuring blood pressure and assessing adherence, and referral for clinical review if needed. The first visit will occur one month after the initial clinic appointment. The frequency of subsequent follow-up visits scheduled based on blood pressure control and adherence status. The full follow-up schedule, including decision points and medication supply intervals, is presented in a flowchart in Figure 1.

**Figure 1.**
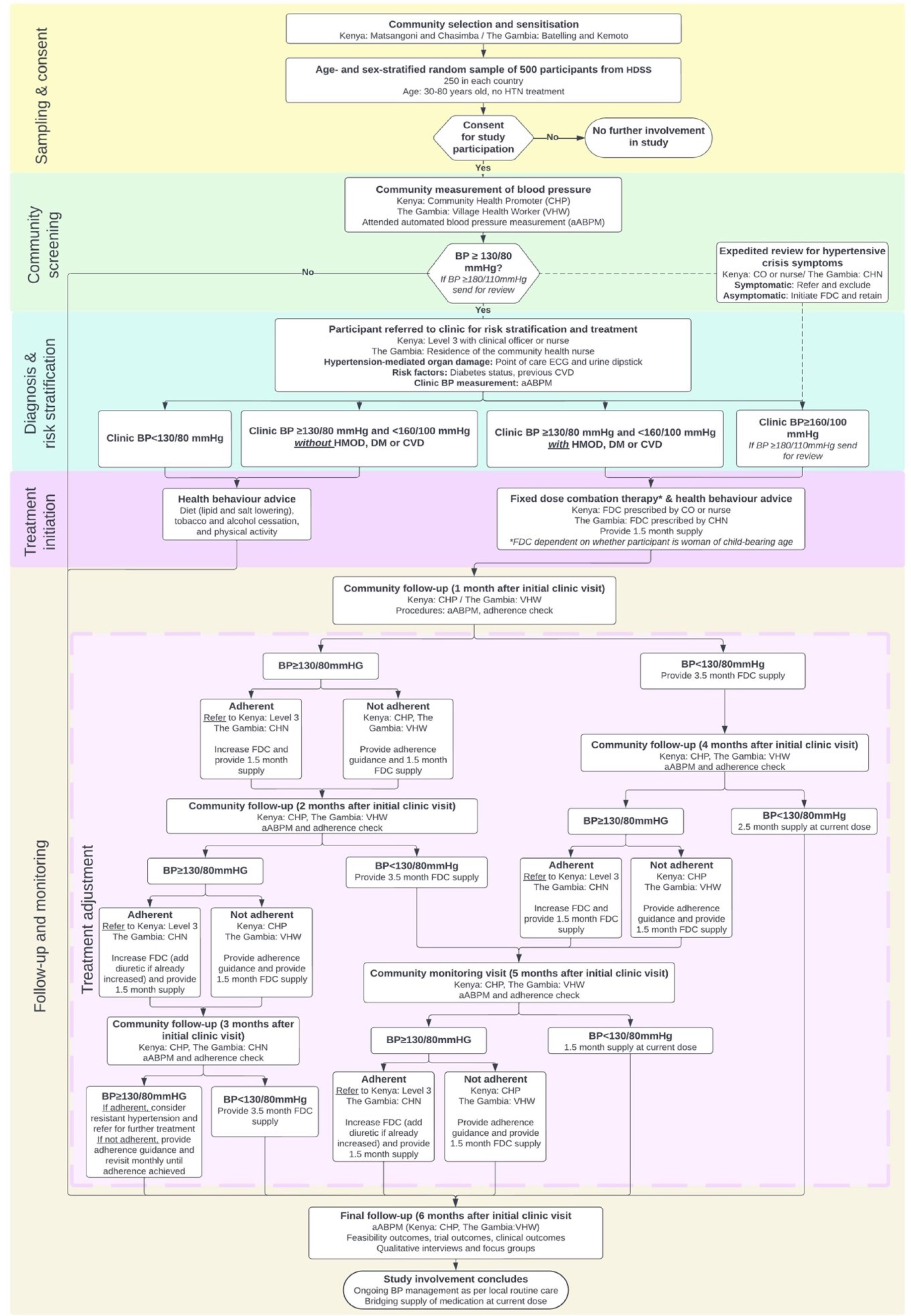
Schematic showing study and clinic activities for all participants involved in the IMPLEMENT-IHCoR feasibility study.

Participants who have controlled blood pressure (<130/80mmHg) will receive medication refills. Those with elevated blood pressure (>130/80mmHg) will have their adherence assessed. If they are non-adherent, they will receive adherence advice and medication refills. If adherent, they will be referred for clinical review and possible dose adjustment. Details of the SPC components and doses are provided in Table 1.

**Table 1.** Details of the SPC components and doses.

| Stage | SPC components and doses |
| --- | --- |
| Initiation of treatment dose - standard | Perindopril (4 mg) + Amlodipine (5 mg) |
| Initiation of treatment dose – women aged 30-49 (childbearing age) | Amlodipine (5 mg) + Hydrochlorothiazide (12.5 mg) |
| Increased dose - standard | Perindopril (8 mg) + Amlodipine (10 mg) |
| Increased dose - women aged 30-49 | Amlodipine (10 mg) + Hydrochlorothiazide (25 mg) |
| Addition of third component - standard | Perindopril (8 mg) + Amlodipine (10 mg) + Hydrochlorothiazide (12.5 mg) |
| Addition of third component - women aged 30-49 | Amlodipine (10mg) + Hydrochlorothiazide (25mg) + Methyldopa (500mg) OR<br>Amlodipine (10mg) + Hydrochlorothiazide (25mg) + labetalol (200mg) |

All participants, including those not on SPC treatment, will receive a final follow-up visit at six months.

Upon study conclusion at 6 months following the initial clinic visit, participants will continue with blood pressure management as per local routine care. A bridging supply of medication will be provided for participants initiated on SPC treatment during the study period.

Participants are permitted to continue any regular medications for existing conditions (e.g., HIV, diabetes, TB). Women who become pregnant during the study will be excluded from the study and referred to usual care. No other specific concomitant care restrictions or allowances are noted in the intervention protocol.

### Training of healthcare providers

Training will be provided to all healthcare providers (CHPs/VHWs, clinical officers, nurses) involved in the delivery of all four components of the intervention (household-level screening, primary healthcare facility risk stratification and diagnosis, initiation of SPC hypertension treatment and/or lifestyle advice, and household-level monitoring and follow-up).

Based on a scoping review of training characteristics conducted by the IMPLEMENT-CVD team, training will be delivered over a maximum of three days, with one day for a practical session. Training will include both lecture-style and interactive components. Training content will be developed and delivered by the study team with support from the Kenya Cardiac Society in Kenya and Cardiac Society of The Gambia. Participants will receive a training booklet and be supported through ongoing mentoring during implementation. Healthcare providers will receive a certificate on training completion, a recommendation that emerged strongly during the co-design workshops.

## OUTCOMES

For Objective 1, we will assess adoption, fidelity, reach, and dose, (defined in Table 2), using a mix of quantitative measures, qualitative methods, and observations.

**Table 2.**
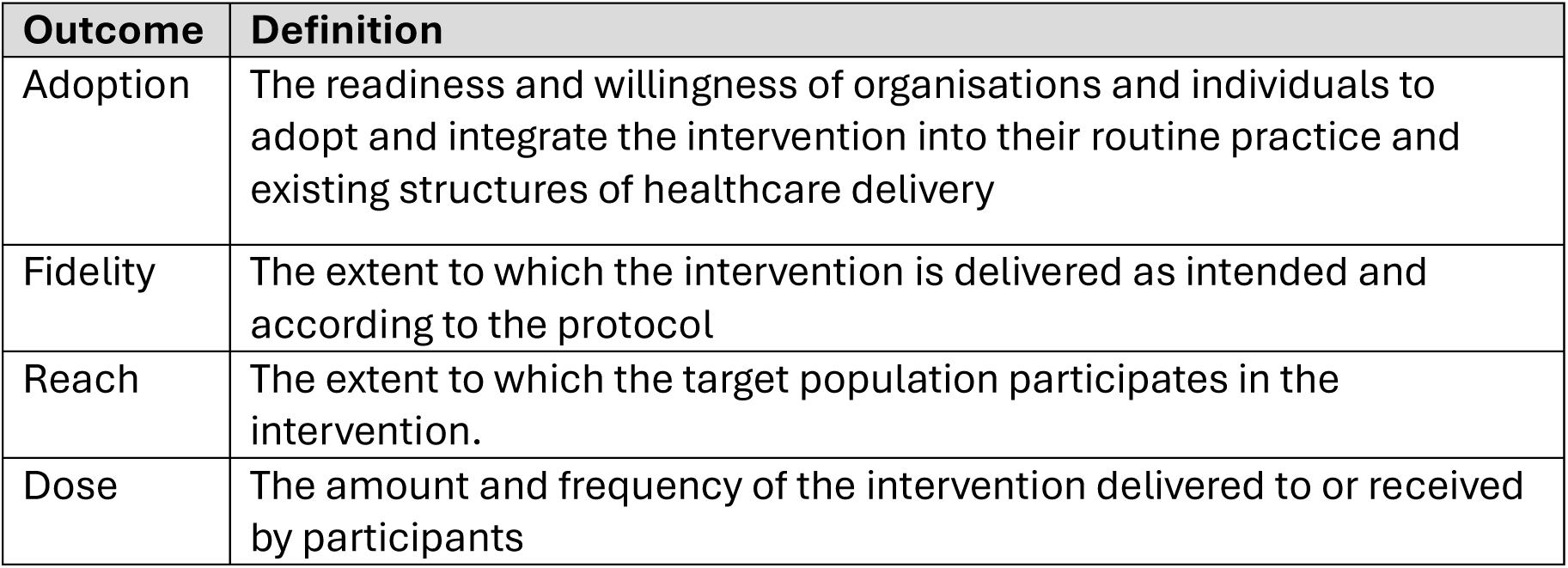
Outcome definitions and assessment for Objective 1.

| **Outcome** | **Definition** |
| --- | --- |
| Adoption | The readiness and willingness of organisations and individuals to adopt and integrate the intervention into their routine practice and existing structures of healthcare delivery |
| Fidelity | The extent to which the intervention is delivered as intended and according to the protocol |
| Reach | The extent to which the target population participates in the intervention. |
| Dose | The amount and frequency of the intervention delivered to or received by participants |

Objective 2 explores mechanisms and contextual factors affecting the feasibility of implementing each intervention component, guided by Normalisation Process Theory [33]. This will involve qualitative interviews and focus group discussions (FGDs) with those delivering the intervention, focusing on:

1. Contextual influences on intervention adoption, fidelity, reach, and dose (as defined in Objective 1).
2. Intervention adaptations due to complexity, participant needs, or contextual constraints.
3. Integration and compatibility of the intervention within existing practices.
4. Political, social, and economic factors affecting implementation and sustainability.

For Objective 3, we will assess intervention acceptability and sustainability from the perspectives of patients, caregivers, healthcare providers, and policymakers through qualitative interviews and FGDs. Acceptability will be evaluated using the Theoretical

Framework of Acceptability, examining seven key constructs: affective attitude, burden, ethicality, intervention coherence, opportunity costs, perceived effectiveness, and self-efficacy [32]. Sustainability will be considered from a broader health system perspective, building on contextual insights and implementation mechanisms explored in Objective 2.

For Objective 4, we will assess costs to the healthcare system and to patients and households. Costs to the healthcare system will include resource use such as staff time, equipment, diagnostics, and overheads, expressed as financial and economic costs. Costs to patients and households, will include direct out-of-pocket expenses and indirect costs such as lost income or time spent seeking care.

The outcomes for Objective 5 are quantitative measures of study methods, including the collection of clinical outcomes, to assess the design of the study and inform the design of future studies [40]. These outcomes and their definitions are given in Table 3.

**Table 3.** Outcome definitions and assessment for Objective 5.

| **Outcome** | **Definition** |
| --- | --- |
| Sampling proportion | Number of participants who were screened for eligibility compared to the number of participants sampled from the HDSS |
| Eligibility proportion | Number of participants eligible compared to the number who were screened. |
| Consenting proportion | Number of participants who gave consent to participate compared to the number of eligible participants. |
| Screening proportion | Combined sampling, eligibility, and consenting rates |
| Data completeness | Percentage of missing data overall and for each case report form in the final dataset. |
| Data accuracy | Percentage of data fields requiring querying and correcting during data cleaning, for each case report form and in total. |
| Proportion lost to follow-up | The percentage of participants who do not attend follow-up visits and do not formally withdraw. |
| Proportion delayed follow-up | The percentage of follow-up visits that are completed outside of the defined window across all visits. |
| Withdrawal ratio | The percentage of participants who do not attend follow-up visits and formally withdraw from the study. |
| Retention ratio | 100% less the combined percentages of loss to follow up and withdrawal ratios at each follow-up timepoint. |
| Blood pressure | Systolic and diastolic blood pressure readings |
| Major adverse CVD events | Self-reported myocardial infarction, stroke, heart failure or hospitalisation for CVD |

## DATA COLLECTION

Data collection methods have been selected to align with the outcomes for each objective. Table 4 shows a summary of objectives, outcomes, and data sources.

**Table 4.** Summary of objectives, outcomes, and data sources.

| **Objectives** | **Outcomes** | **Definition** | **Data source** |
| --- | --- | --- | --- |
| 1. Evaluate feasibility of the intervention | Adoption | Willingness/ readiness to adopt the intervention | Structured interviews and FGDs |
|  | Fidelity | Delivery as intended per protocol | Referral forms, clinic records, and CRFs of clinic and community visits; Structured observations |
|  | Reach | Participation rate among the target population | Screening logs |
|  | Dose | Frequency/amount of intervention delivered or received | Referral forms, clinic records, and case report forms (CRFs) for clinic and community visits |
| 2. Understand mechanisms and contextual factors affecting feasibility | Contextual influences on implementation; adaptations; integration and compatibility with existing health system. | - | Structured interviews and FGDs |
| 3. Understand the acceptability and sustainability of the intervention |  | - | Structured interviews and FGDs |
| 4. Estimate implementation costs | Costs to healthcare system | Resource use, personnel time, overheads, supplies | Facility costing tool; financial/ project records; time and motion observations; staff timesheets |
|  | Costs to patients/ households | Out-of-pocket costs, indirect costs | Patient costing tool |
| 5. Evaluate study procedures and design feasibility | Sampling rate | Screened vs sampled from HDSS | Screening logs |
|  | Eligibility rate | Eligible vs screened | Screening logs |
|  | Consenting rate | Consented vs eligible | Screening logs |
|  | Screening rate | Combined sampling, eligibility, and consent | Screening logs |
|  | Data completeness | % of missing fields per CRF | CRFs; data cleaning logs |
|  | Data accuracy | % of fields requiring correction | CRFs; data cleaning logs |
|  | Loss to follow-up rate | % not attending follow-up & not withdrawn | Clinic records; referral forms |
|  | Delayed follow-up rate | % of follow-ups outside visit window | Clinic records; referral forms |
|  | Withdrawal ratio | % withdrawn from study | Withdrawal logs |
|  | Retention ratio | % retained = 100% – loss/withdrawal | Clinic records, and case report forms (CRFs) for clinic and community visits |
|  | Blood pressure | Systolic and diastolic measurements | Clinic and community BP logs |
|  | Major CVD events | Self-reported events: MI, stroke, etc. | Participant questionnaire |
|  | Medication adherence | Adherence to SPC therapy | Pill counts; MARS-5 questionnaire |

### Objectives 1 and 5 (quantitative data)

For objective 1, implementation outcomes will be measured using:

- Screening logs, completed by CHPs (Kenya) and VHWs (The Gambia), will record participant availability, eligibility, and consent status, informing the measurement of reach.
- Referral forms, clinic records, and case report forms (CRFs) for clinic and community visits to capture whether participants follow the intervention pathway and the extent of intervention delivery, informing measures of fidelity and dose.
- Structured non-participant observations during community and clinic visits to assess fidelity to protocol, including adherence to SOPs and use of intervention tools (discussed further below).

To assess feasibility of the trial design and the quality of clinical outcome data (Objective 5), quantitative data collection will include:

For objective 5, data on trial feasibility will be collected from:

- Screening logs to assess recruitment indicators (sampling, eligibility, and consent rates).
- Clinic records and case report forms (CRFs) for clinic and community visits to evaluate data completeness and accuracy, participant retention, adherence to visit schedules, and withdrawal.
- Blood pressure readings recorded during clinic and community visits.
- Self-reported major CVD events through a questionnaire during follow-up visits.
- Adherence to medication gathered using pill counts and the MARS-5 questionnaire during follow-up visits.

### Objectives 2 and 3 (qualitative data)

Qualitative data collection will be used to support Objective 1, by exploring the mechanisms and contextual factors affecting the feasibility of implementing each intervention component (Objective 2), and the acceptability and sustainability of the intervention (Objective 3).

Semi-structured interviews and focus group discussions (FGDs) will be conducted with patients enrolled in the intervention, caregivers identified by patients, healthcare providers (including CHPs/VHWs) involved in intervention delivery, registered community pharmacists, trainers of healthcare workers, and relevant policy stakeholders at county, regional, and national levels. Purposive sampling will guide participant selection across these groups.

Data collection will be conducted in participants’ preferred language by trained staff. Topic guides will be informed by the Theoretical Framework of Acceptability and Normalisation Process Theory. Regular team debriefs will inform iterative refinement.

Two patient groups will be included: those receiving lifestyle advice only (via FGDs post-intervention or at study end), and those initiated on treatment (interviewed after the one-month follow-up and again at months 4–5). A subset of patients who miss appointments will be purposively selected to reflect a range of intervention experiences. Caregiver interviews will be conducted during months 3–5 and may occur jointly with patients, with selection based on gender, age, and relationship to the patient.

Healthcare providers will be interviewed post-treatment initiation and again at months 4–6 to assess training, diagnosis, lifestyle advice, and treatment. CHPs/VHWs will be interviewed following their first follow-up visit and again at months 4–6 to capture screening and monitoring experiences. Community pharmacists will be interviewed in months 3–6 to explore their training experiences and interactions with patients receiving SPC treatment.

Trainers of healthcare providers will be interviewed post-training covering training content, delivery, mentoring experience, and recommendations for improvement.

County, regional, and national policy stakeholders will be interviewed from 3month onward to assess views on implementation, health system implications, and the potential for scale-up and sustainability.

### Objective 4 (costing data)

For objective 4, costing data will be collected using two standardised tools:

- The facility costing tool, which records inputs such as personnel time, diagnostics, medications, and facility overheads, enabling bottom-up and top-down cost estimates from the provider perspective.
- The patient costing tool, which captures direct medical (e.g., treatment costs), direct non-medical (e.g., transport, food) out-of-pocket costs and indirect/productivity costs from participants.

These tools will be completed through structured interviews with healthcare providers and participants, in addition to a review of financial and project administrative records.

Prospective time and motion studies will be conducted through direct observations (discussed further below), and self-reported timesheets.

### Observations to support Objectives 1 and 4

Structured non-participant observations will be conducted to support the assessment of fidelity to intervention protocols during screening, clinical assessment, and follow-up (Objective 1) and for the time and motion studies (Objective 4).

Observations will include:

- CHPs (Kenya) or VHWs (The Gambia) performing community-based aABPM.
- Clinical officers or nurses conducting clinic-based assessments (ECG, urine dipstick, and treatment provision).

All observations will occur during routine intervention delivery using a standardised checklist aligned with SOPs. The checklist will capture key fidelity indicators such as adherence to protocol steps, appropriate tool usage, and provider-participant interactions. Observational data will be recorded directly onto the checklists.

## SAMPLE SIZE

### Objectives 1 and 5 (quantitative data)

We will screen 500 participants (250 in Kenya and 250 in The Gambia). The sample size was determined based on feasibility considerations (time, budget, and recruitment capacity) and to allow sufficiently precise estimates of key implementation and trial design outcomes, in line with published guidance [27, 40]. The IMPLEMENT-IHCoR intervention includes blood pressure screening, diagnosis, CVD risk stratification, treatment, and follow-up components. Therefore, the number of participants receiving the intervention varies by component (see Figure 1). Based on prior IHCoR-Africa data and the planned intervention design, we anticipate that 45% (n=225) of participants will be referred from community screening to clinic for diagnosis and risk stratification. Of these, we estimate that 25% will be recommended for SPC therapy. In total, we estimate that just over 10% (55 participants) of the total study sample will be recommended for SPC therapy.

Precision estimates for key outcomes were calculated using these assumptions. For example, with an expected clinic attendance rate of 90%, we would achieve ±5% precision (95% CI: 85.1%–93.4%). Table 5 presents confidence intervals for screening reach, clinic attendance, treatment adherence, and retention across relevant subgroups.

**Table 5.** Precision estimates for key outcomes based on a sample size of 500 participants.

| Outcome | Number of participants providing data | Assumed proportion achieving outcome threshold | 95% confidence intervals |
| --- | --- | --- | --- |
| Community blood pressure assessment (reach) | 500 | 95.0% | 92.7% - 96.7% |
| Clinic HMOD and risk factor assessment (reach) | 225 | 90.0% | 85.1% - 93.4% |
| Treatment adherence (dose) | 55 | 80% | 67.0% - 89.5% |
| Retention rate (dose) | 500 | 80% | 76.2% - 83.4% |

### Objectives 2 and 3 (qualitative research)

Sample size will be determined based on participant eligibility, team capacity, participant burden, and need for meaningful site comparisons. Data collection will occur at two key timepoints: approximately one month after initial intervention delivery and again at four to five months to explore perceptions over time. Anticipated interview numbers per group and site are outlined in Table 6.

**Table 6.** Sample size by group, for each study site.

| **Data collection method** | **Participant category** | **Kenya** | **The Gambia** |
| --- | --- | --- | --- |
| In-depth interview | Patients screened, referred & initiated on medication | 15 | 15 |
|  | Family caregivers of patients screened, referred & initiated on medication | 7 | 5 |
|  | CHPs/VHWs | 10 | 3 |
|  | Healthcare providers | 10 | 7 |
|  | Trainers/mentors of healthcare workers | 5 | 3 |
|  | Community pharmacists | 2 | 0 |
|  | Policy stakeholders | 8 | 5 |
|  | Total number of interview participants | 57 | 38 |
| FGD | Study participants screened, referred, not initiated on medication | 4 FGD with 6-8 participants | 2 FGD with 6-8 participants |
|  | Total number of FGDs | 24-32 | 12-16 |
| Observations | CHPs/VHWs | 10 CHPs (5 from each CHU), each observed twice | 3 VHWs, each observed twice |
|  | Healthcare providers (Clinical Officers/Nurses) | 4 clinical officers, each observed twice | 4 CHNs, each observed twice |
|  | Total observation records | 28 | 14 |

### Observations to support Objectives 1 (fidelity) and 4 (time motion study)

In Kilifi, Kenya, the Matsangoni and Chasimba CHUs include 2 clinical officers, one in each site, and 38 CHPs (12 and 26, respectively). All clinical officers (n=2) will be observed. A random sample of 10 CHPs (5 per site) will be selected, with final sample size adjusted based on observed variability, intervention complexity, and logistical feasibility. In Kiang West, The Gambia, observations will include all 3 VHWs and 4 CHNs across the study villages of Batelling, Kemoto, and Tankular. Numbers per group and site are outlined in Table 6.

## DATA MANAGEMENT

All study data will be securely stored and managed in compliance with local and international regulations. Hard copies (e.g. consent forms) will be stored in locked cabinets at each study site. Electronic data will be encrypted and stored on secure servers at KWTRP, MRCG, and LSHTM, with access restricted to authorised personnel through multi-factor authentication and role-based access controls. Data transfers between institutions will use encrypted, password-protected platforms, governed by existing data-sharing and collaboration agreements. All individual records will be de-identified and indexed using unique study identifiers, and no names or directly identifiable information will be retained. De-identified study materials will be archived at each partner institution for a minimum of 10 years, in line with institutional policies. A comprehensive data dictionary and standard operating procedures (SOPs) will be developed to guide data handling and ensure consistency across sites and teams.

### Quantitative and costing data

Quantitative and costing data will be collected electronically by trained study staff using REDCap on password-protected Android tablets. Case report forms (CRFs) will be programmed with built-in validation logic to reduce entry errors and improve real-time data quality. Tablets will be wiped after data transfer. Uploaded data will reside on encrypted servers hosted by KWTRP and MRCG. Data discrepancies or inconsistencies will be addressed systematically through data queries. Cleaned datasets will be securely stored and retained at LSHTM, KWTRP, and MRCG.

### Qualitative data

Qualitative data from interviews and focus group discussions will be audio-recorded and transcribed verbatim. In Kenya, transcripts in Kiswahili or Giriama will first be transcribed in the source language and then translated into English. In The Gambia, recordings in Mandinka will be simultaneously translated and transcribed into English. Transcripts will be reviewed for accuracy and de-identified, with names and locations replaced by descriptors (e.g., “nurse”, “health facility”). Transcribed data will be imported into NVivo 12 for thematic analysis.

## DATA ANALYSIS

### Quantitative data analysis (Objectives 1 and 5)

Descriptive statistics (means, proportions, and confidence intervals) will be used to summarise participation rates, referral uptake, and intervention delivery frequency, and study design metrics (sampling, eligibility, consent, data completeness, loss to follow-up, and retention). Data will be provided by screening and follow-up logs, referral forms, and electronic CRFs. Structured observation checklists will be used to assess fidelity by calculating adherence rates to SOP-defined procedures. Data analysis will be conducted using the statistical software packages R and Stata.

### Qualitative data analysis (Objectives 2 and 3)

Qualitative data will be analysed thematically using both inductive and deductive approaches. Three coding frameworks will be developed: (1) patients/caregivers, (2) healthcare providers/CHPs/community pharmacists, and (3) policymakers/decision-makers.

Two analysts will review all transcripts to familiarise themselves with the data and conduct line-by-line open coding on five diverse transcripts. The analysis team will meet to compare initial codes and produce a shared framework of concepts, relationships, and practices. This framework will be applied to an additional five transcripts, refined through discussion, and then applied to the remaining data. Weekly team meetings will support ongoing refinement.

Coded data will be mapped to constructs from both the Theoretical Framework of Acceptability (TFA) and Normalisation Process Theory (NPT). Comparative analysis will be conducted across datasets and constructs. Findings will be synthesised into a narrative reflecting contextual influences, mechanisms of implementation, and factors shaping acceptability and sustainability. NVivo 12 will be used to manage all qualitative data.

### Cost Analysis (Objective 4)

To ensure contextual relevance, we will prioritize the use of local price data from Kenya and The Gambia to value both traded and non-traded goods. Where local price data is unavailable, national averages or regional estimates from comparable settings will be used. International prices will be applied to traded goods in one-way sensitivity analyses, adjusted using purchasing power parity (PPP) or market exchange rates, as appropriate [41]. All goods and services will be clearly classified as traded or non-traded in the dataset to support transparency and enhance transferability of findings to other settings.

Where market prices are not available—such as for donated goods or community health provider (CHP) time—we will estimate shadow prices. These will be based on the average of multiple local market estimates or, where necessary, derived from the opportunity cost (e.g., the wage a CHP could earn based on their qualifications) or the replacement cost (e.g., the wage of a typical worker performing similar tasks).

We will calculate total cost and per-patient costs across three distinct implementation phases: (1) pre-implementation (planning and start-up), (2) implementation (screening, risk stratification, and treatment), and (3) full implementation (ongoing delivery).

Costs will be categorized as capital or recurrent, and as economic or financial. Capital costs will be annuitized over the expected useful life using a 3% discount rate. Financial costs will be depreciated using straight-line methods. Local estimates of useful life will be used where available; where not, standard values from comparable settings will be applied. One-way sensitivity analyses will be conducted on key cost parameters. Where feasible, probabilistic sensitivity analysis will also be performed to assess the robustness of cost estimates.

## DATA MONITORING

A formal data monitoring committee was not established, as the study involves minimal risk and does not include a control arm or therapeutic randomisation. Safety will be assessed and monitored by the study staff on an ongoing basis. The study team is well-equipped to oversee participant safety through regular and systematic assessments and monitoring of adverse events and emerging safety concerns. Any adverse effects will prompt immediate clinical review and, if needed, withdrawal from the study. No external auditing is planned. Internal oversight will be conducted by the study management team.

## ETHICS APPROVAL

### Ethics approval

Ethics approval was obtained from the Kenyan National Committee for Science, Technology and Innovation (NACOSTI) (ref: 415561), the Gambia Government/Medical Research Council Joint Ethics Committee (ref: 31372) and the London School of Hygiene and Tropical Medicine Ethics Committee (ref: 31372).

### Informed consent

Informed consent will be obtained from all the study participants prior to any study activities commencing. CHPs and CHNs/VHWs will receive detailed training on the consent procedures and will be responsible for obtaining consent. We will put in place all necessary measures to remove barriers to providing informed consent and participating in the research, such as language and literacy.

All study participants will be provided with comprehensive information about the study in the language that they are most comfortable with. This information will explain the aims of the study, their specific role in the study, and how data that is being collected will be used. Where a participant is illiterate an independent witness will be present to ensure the correct study information is presented to the participant. This witness will also sign the consent form. It will be made clear that participation in any part of the research will be entirely voluntary, will not influence future care and that participants will be free to withdraw at any stage, without giving a reason.

### Risks

During the study, it possible that severe uncontrolled hypertension could be identified. In such cases we will facilitate referral to urgent care for participants. Further treatment will follow the locally available standard of care. Medications can cause side effects for some patients. If prescribed medications are causing significant side effects, alternative medications will be prescribed.

### Ancillary and post-trial care

Participants who are diagnosed with hypertension during the study will be referred to local health services for continued care following study completion. A bridging supply of hypertension medication will be provided at study end. Any incidental health concerns identified during the study will be communicated to participants and referred to appropriate healthcare providers.

## DISSEMINATION

Study findings will be disseminated through multiple channels to reach diverse audiences, including academic, policy, and community stakeholders. Results will be shared via peer-reviewed journal publications, national and international conferences, and policy briefs targeted at health system decision-makers. Lay summaries will be shared through our well-established quarterly IHCoR newsletter and partner newsletters. Dissemination of study findings to participants and the wider community will be done through our community engagement forums which we established in earlier phases of this project.

## CONCLUSION

This study addresses a critical public health challenge of the high burden of undiagnosed and poorly managed hypertension in rural SSA. By evaluating the feasibility of a community-based approach that leverages community health workers to identify, risk-stratify, and manage individuals with hypertension, this study aims to promote earlier diagnosis and timely care, potentially preventing serious cardiovascular complications. The findings will generate novel evidence on the implementation of integrated, task-shared hypertension care in rural SSA. This will inform the design of a future full-scale trial and contribute to the development of scalable, cost-effective strategies to strengthen hypertension management and improve health outcomes in similar contexts.

## STUDY STATUS AT TIME OF SUBMISSION

At the time of protocol submission (August 4, 2025), recruitment was completed in both Kenya and The Gambia (250 participants were screened per country). In The Gambia, 107 participants were referred to the clinic. Of those, 17 were initiated on SPC therapy and 80 received health behaviour advice only. Two patients were not yet seen at the clinic, 4 were lost to follow-up, and 1 withdrew from the study. In Matsangoni, 73 participants were referred to clinic. Of those, 18 were initiated on SPC therapy and 45 received health behaviour advice only, and 10 were lost to follow-up. In Chasimba, 78 participants were referred to clinic. Of those, 24 were initiated on SPC therapy and 39 received health behaviour advice only, while 15 participants have not yet been seen at clinic. First month follow-up visits are currently underway in both countries.

## AUTHOR CONTRIBUTIONS

- Conception or design of the work – SH, NK, BD, RW, JH, RW, AB, CK, CM, AS, AP, NM, EN, BT, EB, PP, AOE, MJ, AM
- Drafting the article – SH, NK
- Critical revision of the article – SH, NK, BD, RW, JH, RW, AB, CK, CM, AS, AP, NM, EN, BT, EB, PP, AOE, MJ, AM
- Final approval of the version to be submitted – SH, NK, BD, RW, JH, RW, AB, CK, CM, AS, AP, NM, EN, BT, EB, PP, AOE, MJ, AM

## DATA AVAILABILITY STATEMENT

This is a protocol and there is no data underlying this article.

## ACKNOWLEDGEMENTS

IHCoR-Africa Collaborators: Andrew Prentice, Assan Jaye, Abba Hydara, Bai Cham, Samson Kinyanjui, Elijah Ogola, Jemima Kamano, Violet Naanyu, Lilian Mbau, Tim Clayton, Melanie Morris, David Prieto-Merino, Cova Bascaran, James Abuje, Mavis Foster-Nyarko, Saidina Ceesay, Ruth Lucinde, David Leon, Emily Herret.

